# Use of the EsoGuard^®^ Molecular Biomarker Test in Non-Endoscopic Detection of Barrett’s Esophagus among High-Risk Individuals in a Screening Population

**DOI:** 10.1101/2024.06.24.24309401

**Authors:** Nicholas J. Shaheen, Mohamed O. Othman, Jawar Taunk, Kenneth J. Chang, Sathya Jaganmohan, Patrick S. Yachimski, John C. Fang, Joseph S. Spataro, Suman Verma, Victoria T. Lee, Brian J. deGuzman, Lishan Aklog

## Abstract

**Background and Aims:** Barrett’s Esophagus (BE) is the precursor to esophageal adenocarcinoma (EAC). We aimed to assess performance, safety, and tolerability of the EsoGuard (EG) assay on samples collected non-endoscopically with the EsoCheck (EC) device (EG/EC) for BE detection in the intended-use population, meeting American College of Gastroenterology (ACG) guideline criteria (chronic gastroesophageal reflux disease (GERD) and 3+ additional risk factors).

**Methods:** We performed a prospective, multicenter study (NCT04293458) to assess EG performance (primary endpoint) on cells collected with EC, for detection of BE and EAC using esophagogastroduodenoscopy (EGD) and biopsies as the comparator. Twenty-four sites across the U.S. and Spain participated. EC safety and usability were assessed as secondary endpoints.

**Results:** 180 male subjects aged >50 years with chronic GERD met eligibility criteria, of which 163 (90.6%) had EGD and successful EC administration. Mean age was 60.5yrs, 34.4% were obese, 56.7% had tobacco history, and 3.9% had a 1^st^ degree relative with BE or EAC. Of 122 samples analyzed, 93 contributed to the primary endpoint analysis. About 9% of subjects in the Primary Analysis Population had BE on EGD, none with dysplasia. Sensitivity of EG for BE was 87.5% (95% CI 47.4-99.7), specificity was 81.2% (95% CI 71.2-88.8), positive predictive value was 30.4% (95% CI 13.2-52.9), and negative predictive value was 98.6% (95% CI 92.3-99.96). Mild esophageal abrasions were observed in 1.5%; no serious adverse events were reported.

**Conclusions:** EG/EC appears effective for BE screening. This approach provides a safe, accurate, and well-tolerated non-endoscopic alternative in high-risk patients.

## Introduction

Barrett’s Esophagus (BE) is the only known precursor to esophageal adenocarcinoma (EAC), the most common esophageal cancer in Western countries, with an increasing incidence over the last several decades.[1–4] BE is a metaplastic condition of the esophagus with well-established risk factors, including chronic gastroesophageal reflux disease (GERD).[5] In contrast to the lethality of EAC, with a 5-year mortality of ∼80%, BE can be successfully treated using endoscopic strategies which achieve complete disease eradication in ≥80% of patients.[6–10]

Endoscopic screening and surveillance of BE is supported by multiple gastroenterology societies, with the goal of reducing incidence of EAC and EAC-related mortality.[11–13] The American College of Gastroenterology (ACG) recommends screening patients with chronic GERD and ≥3 of the following risk factors: male sex, White race, age >50 years, tobacco smoking, obesity, and family history of BE or EAC in a first degree relative.[11] Unfortunately, literature shows only 10-30% of eligible patients with chronic GERD undergo endoscopic BE screening.[14–16] Instead, diagnoses of BE are most commonly made when patients with refractory or severe GERD symptoms undergo esophagogastroduodenoscopy (EGD); however many GERD patients are on acid suppressive medications with good symptom control.[17, 18] These patients may not be referred by primary care providers for screening EGDs, resulting in missed BE diagnoses. Additionally, most BE risk factors (i.e., male sex, age >50 years, white race) are highly prevalent; with GERD seen in up to 44% of many Western populations,[19] and it is impractical to perform EGD on all individuals meeting guideline criteria, due to resource limitations. This clinical gap is well-recognized, and the latest ACG guidelines suggest non-endoscopic modalities as a reasonable alternative to EGD for initial BE screening.[11]

EsoGuard® (EG) is a commercially available test for BE, performed in a Clinical Laboratory Improvement Amendment (CLIA) certified lab (LucidDx Labs, Lake Forest, CA). EG utilizes methylated DNA biomarkers to detect BE and is performed on cells collected non-endoscopically with EsoCheck® (EC), an FDA 510(k) cleared device for collection of mucosal cells in the esophagus. Performance characteristics of EG on cells collected with EC (EG/EC) have previously been described in two case-control studies, demonstrating excellent sensitivity and specificity for detection of both BE and EAC.[20, 21] Here we present the first results of EG/EC for BE screening in the intended-use population of individuals meeting ACG guideline criteria. We also describe alterations in specimen processing to increase feasibility of implementing EG/EC as a point-of-service test.

## Methods

### Study Design and Participants

We performed a prospective, multi-center, single-arm study recruiting subjects from a BE screening population, defined as endoscopy-naïve individuals warranting BE screening according to ACG’s guidelines. The primary objective was to assess performance of EG for detection of BE in this population. EGD plus biopsies to establish a histopathologic diagnosis was the diagnostic comparator; all subjects underwent cell collection first with EC, and EGD second (NCT04293458). EG analysis of cell samples occurred in a delayed fashion, as described below.

Because research staff lacked previous experience using EC, the study consisted of a lead-in EC device administration Training Phase, followed by the Main Study Phase. During the Training Phase, EC device administrators trained in the technical steps and performed a minimum of five proctored collections. Only after completing the Training Phase were sites released to recruit subjects into the Main Study Phase, from which EG/EC performance data were obtained for endpoint analysis. Unlike Main Study Phase subjects, Training Phase subjects did not necessarily meet risk criteria for BE screening; they did not undergo EGD nor were their cell samples analyzed with EG. Safety data were collected for all subjects who underwent EC, whether in the Training or Main Study Phase. Training Phase subjects received final safety follow-up 24-72 hours following EC, and Main Study Phase subjects received final safety follow-up 24-72 hours following EGD. The study was conducted according to the guidelines of the Declaration of Helsinki and approved by local Institutional Review Boards/Ethics Committees for each participating site. All subjects signed informed consent prior to study procedures.

Subject recruitment occurred at 21 U.S. and three Spanish sites. Inclusion criteria required subjects to be male, >50 years old, have a history of chronic GERD defined as ≥5 years of symptoms and/or proton pump inhibitor therapy, and one or more additional BE/EAC risk factors, consistent with ACG guidelines at the time of the study. Details of the eligibility criteria are in **Supplemental Table 1**.

### EsoCheck and EsoGuard

EsoCheck (Lucid Diagnostics, New York, NY) is an FDA 510(k)-cleared, non-endoscopic, encapsulated balloon device designed for circumferential, targeted collection and protected retrieval of cells from the esophagus. EC can be performed in any office setting without sedation, the steps of which are highlighted in **Figure 1**. Controlled device withdrawal across the distal five centimeters of the esophagus allows the textured surface of the inflated EC balloon to collect cells from this target region. It is then deflated and inverted into the capsule, protecting the sample during removal. Outside the body, the balloon is partially re-inflated and cut from the capsule into a preservative solution and transported at ambient temperature to the laboratory.

**Figure 1.**
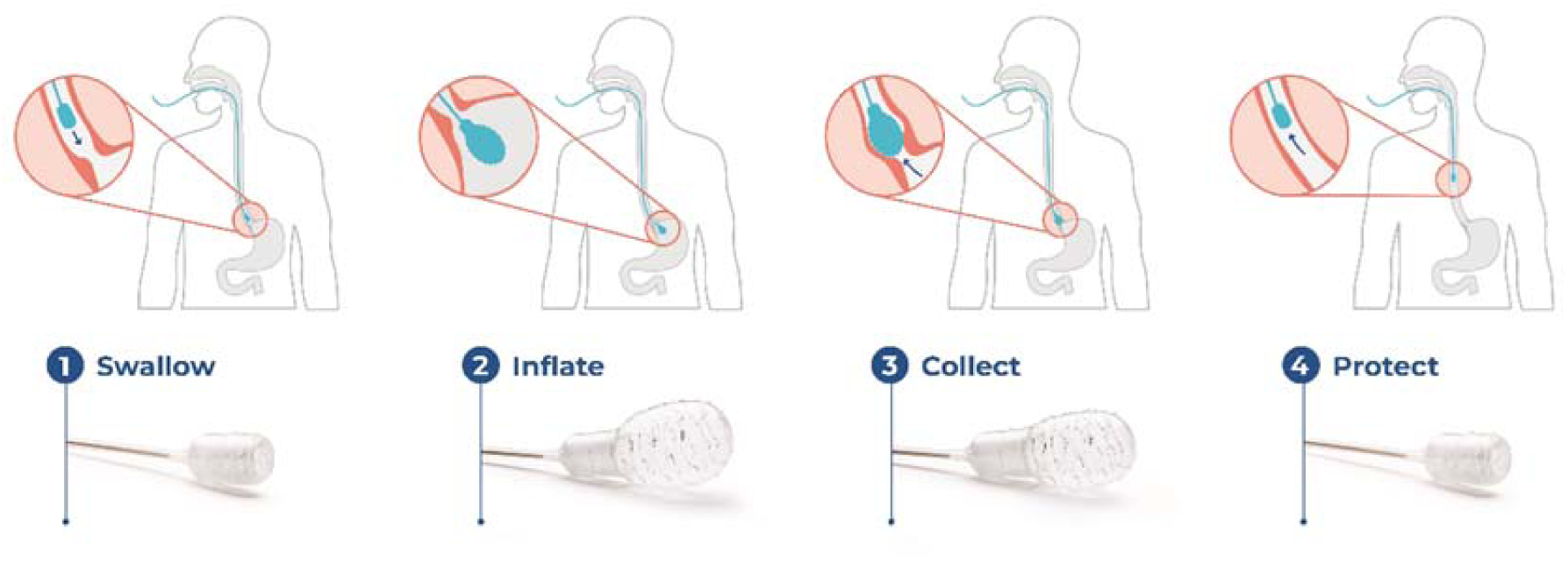
EsoCheck cell collection process.

EsoGuard is a targeted Next Generation Sequencing (NGS) assay assessing methylation on the vimentin (VIM) and Cyclin-A1 (CCNA1) genes. To process specimens, DNA is first extracted, and quantity measured; samples with < 5ng/μl DNA concentration are reported as Quantity Not Sufficient (QNS) for EG analysis. Samples with sufficient DNA undergo bisulfite conversion, polymerase chain reaction (PCR) amplification, library preparation, followed by NGS. Sequencing data are analyzed using a proprietary algorithm and samples passing all quality check (QC) criteria are reported in a binary fashion (positive or negative) indicating whether sufficiently abnormal methylation is present to suggest BE/EAC based on a pre-established cut-off. Any samples failing QC are reported “unevaluable.”

Study samples initially underwent DNA extraction at a partner laboratory (PacificDx, Irvine, CA) within 14 days of receipt and then frozen. They were then transported to LucidDx Labs, with plans to run the EG assay after completion of enrollment. After study initiation, formulation of the preservation media was updated based on internal research indicating the modifications improved methylation signal preservation during sample transport, while allowing for room temperature storage, to improve feasibility as a point of service test. Given these changes, study samples collected in the obsolete version of the transport media (n=34) were not analyzed with EG and were excluded from the performance endpoint analysis.

### EGD, Biopsies, and Diagnosis

On the same day following EC or up to four weeks after, subjects underwent a standard-of-care EGD. In subjects with columnar-lined mucosa of <1cm length (including irregular Z-lines), at least four biopsies were acquired from the area. For subjects with ≥1 cm salmon-colored mucosa on white light endoscopy, the Seattle protocol was followed.[22] Biopsies were not taken of normal-appearing Z-lines. Subjects undergoing EC and EGD on the same day (n=130) underwent visual assessment of the esophageal mucosa for injury, and abrasions were reported as adverse events (AEs). Abrasions were graded for severity according to a previously developed, five-point scale (**Supplemental Table 2**).[23]

Biopsies were interpreted by pathologists at the study sites, then independently adjudicated by expert gastrointestinal pathologist(s) at a central facility (Cleveland Clinic Labs, Cleveland OH). Based on the subjects’ endoscopic and histologic diagnoses, they were assigned to one of 5 groups: 1) ‘negative’ (no specialized intestinal metaplasia/SIM), 2) SIM of the esophagogastric junction (SIM-EGJ, defined as < 1cm cephalad displacement of the Z-line on EGD, with SIM on biopsies), 3) non-dysplastic BE (NDBE, ≥ 1cm of salmon-colored mucosa in the tubular esophagus and SIM without dysplasia), 4) dysplastic BE (SIM with low-grade or high-grade dysplasia) or 5) EAC. If the site and central pathologist disagreed on the diagnosis, a second central pathologist adjudicated the case, with a consensus reading assigned based on the agreement of 2 of the 3 pathologists. For EG performance assessment, study diagnoses were given binary categorization: ‘positive’ denoted presence of BE or EAC on EGD and biopsies; ‘negative’ denoted either normal EGD, or salmon-colored mucosa on EGD *without* SIM on biopsies. Subjects with SIM-EGJ were “indefinite” for BE/EAC and excluded from endpoint analysis.

### Outcome Measures

Performance for detecting BE/EAC (primary endpoint) was assessed using (1) sensitivity; (2) specificity; (3) positive predictive value (PPV); and (4) negative predictive value (NPV). EC safety (secondary endpoint) was based on rates of AEs deemed by the site investigator as possibly, probably, or definitely related to EC. Additional endpoints included EC tolerability, calculated as the proportion of Main Study Phase subjects who successfully swallowed the device and provided a cell sample, and acceptability.

EC acceptability was assessed for Main Study Phase participants, whether successful or not. Subjects completed a two-part survey conducted prior to, and immediately following cell collection, assessing overall experience. A third survey was conducted during the final safety follow-up comparing the EC with EGD experience. Administering personnel completed questionnaires following each cell collection, soliciting satisfaction with the procedure and their assessment of subject tolerance.

### Statistical Analysis

Descriptive statistics were summarized for demographic/baseline characteristics, and relevant medical history. The Main EsoCheck Population (purple box, **Figure 2**) includes Main Study Phase subjects who met all eligibility criteria and underwent EC. The Primary Analysis Population (green box, **Figure 2**) for assessment of EG/EC performance included only Main Study Phase Subjects with binary (i.e., positive, or negative) EG results and binary EGD biopsy-based diagnosis for BE/EAC. Variables collected during EC cell collection and EGD were reported as counts and percentages. EG sensitivity, specificity, and the PPV and NPV for the observed disease prevalence were reported with one-sided asymptotic 95% confidence intervals for binomial proportion without continuity correction. Safety analyses included participants in both the Training and Main Study Phases in whom EC was attempted, whether successful or unsuccessful. Safety events are summarized according to frequency of subjects reporting adverse device-related events (ADEs).

**Figure 2.**
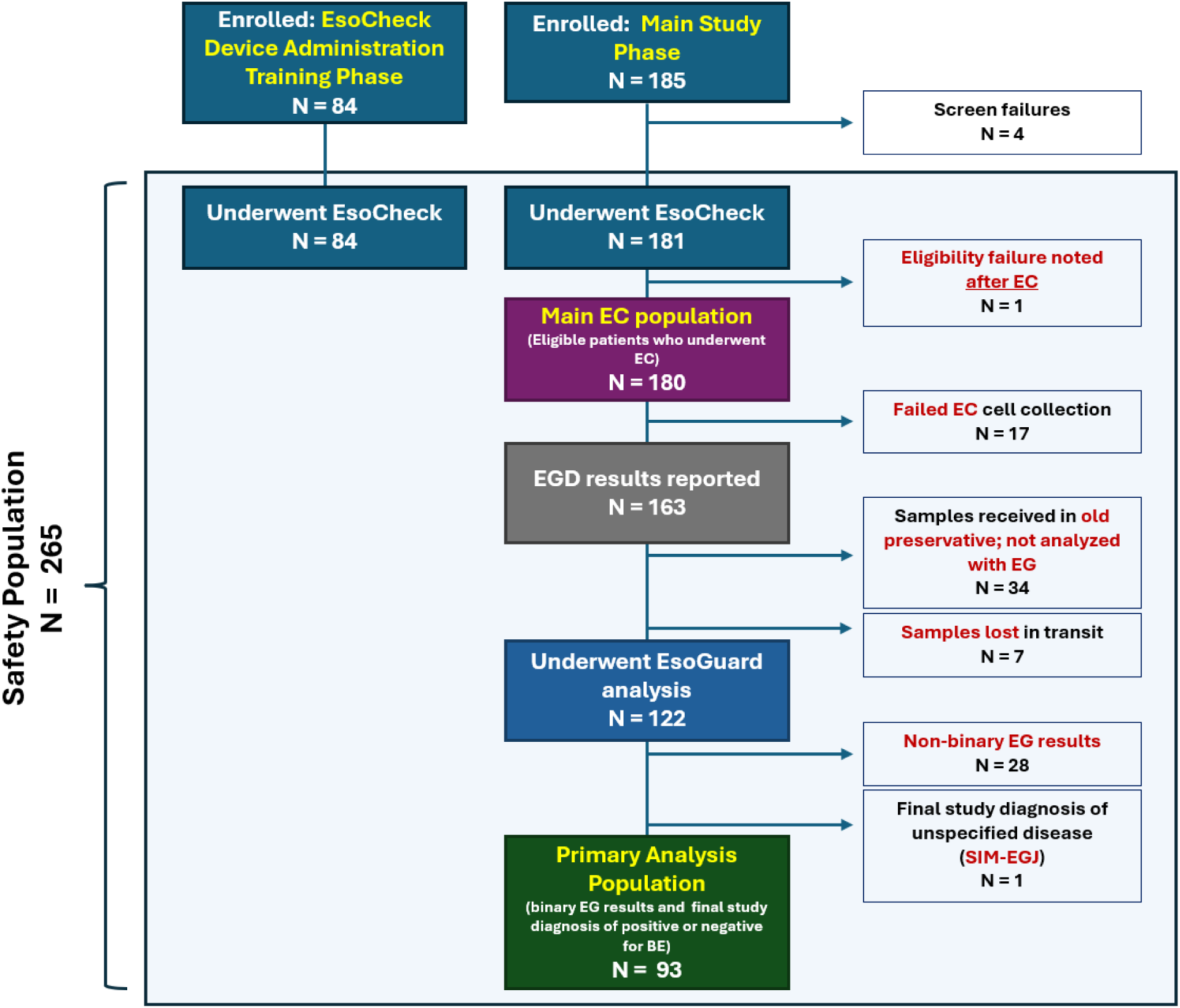
Subject Disposition and Flow of Subjects Through the Study. (EC = EsoCheck; EG = EsoGuard; EGD = esophagogastroduodenoscopy, SIM-EGJ = specialized intestinal metaplasia of the esophagogastric junction, Final study diagnosis = based on EGD and biopsy findings).

## Results

The disposition of study participants is in **Figure 2**. There were 265 subjects from both the EC Training Phase and Main Study Phase contributing safety data. One hundred and eighty-five subjects signed consent for the Main Study Phase, however five failed to meet all eligibility criteria, of which one received EC prior to this discovery and thus still contributed to the safety analysis. The remaining 180 subjects constituted the Main EsoCheck Population. Of these, 17 failed EC and exited the study early; EGD findings were reported for the remaining 163 subjects. Of these, 34 were enrolled prior to implementation of the new preservative and their samples were not analyzed with EG. Seven samples were lost in transit between PacificDx and LucidDx Labs. Thus, 122 samples underwent EG analysis, among which 26 had QNS, two failed QC, and one subject was indeterminate for BE (SIM-EGJ), leaving 93 subjects in the Primary Analysis Population.

Demographic and baseline characteristics of the Main EsoCheck Population are provided in **Table 1**. Mean age was 60.5 ±7.8 years and 54 (30%) were aged > 65 years; 176 (97.8%) were White; 62 (34.4%) obese; 102 (56.7%) had tobacco smoking history, and 7 (3.9%) had family history of BE or EAC in a first degree relative.

**Table 1.** Summary of demographic and baseline characteristics (Main EsoCheck Population)

| **Demographic and baseline characteristics** | **Total (n=180)** |
| --- | --- |
| Age, years |  |
| Mean (SD) | 60.5 (7.8) |
| Median [Q1 - Q3] | 60 [53.0 – 66.5] |
| Min, Max | 50.0, 79.0 |
| Race | **n (%)** |
| White | 176 (97.8) |
| Black | 4 (2.2) |
| American Indian or Alaska native | 0 (0.0) |
| Asian | 0 (0.0) |
| Native Hawaiian or other Pacific Islander | 0 (0.0) |
| BMI (kg/m<sup>2</sup>) |  |
| Mean (SD) | 29.3 (4.7) |
| Median [Q1 - Q3] | 28.6 [26.4 – 31.9] |
| Min, Max | 20.1, 45.8 |
| BMI range (obesity status) | **n (%)** |
| <30 kg/m<sup>2</sup> (non-obese) | 118 (65.6) |
| ≥30 kg/m<sup>2</sup> (obese) | 62 (34.4) |
| Chronic Smoker (past or present) | **n (%)** |
| No | 78 (43.3) |
| Yes | 102 (56.7) |
| Number of years of GERD |  |
| Mean (SD) | 13.9 (10.4) |
| Median [Q1 - Q3] | 10.0 [6.0 – 20.0] |
| Min, Max | 5.0, 57.0 |
| First degree family member with known diagnosis of BE and EAC | **n (%)** |
| No | 173 (96.1) |
| Yes | 7 (3.9) |
| Diagnosis details of family member |  |
| Unknown | 2 (28.6) |
| Esophageal adenocarcinoma (EAC) | 5 (71.4) |

SD = Standard Deviation, Q1 = First quartile, Q3 = Third quartile. Percentages are based on Enrolled Population, N in header.

**Table 2** summarizes the EC and EGD procedures. Median EC cell collection time was four minutes; an outlier (70 min) impacted the mean (5.8 min). Among 163 subjects who contributed EGD results, 40 (24.5%) had findings of salmon-colored mucosa in the tubular esophagus, 20 of which met endoscopic criteria for BE. EGD diagnoses are in **Table 3** for both the Main EsoCheck Population and the Primary Analysis Population. In the Main EsoCheck Population, 13 (7.2%) were positive for BE (all non-dysplastic), 145 (80.6%) were negative; four (2.2%) were SIM-EGJ, and one (0.7%) lacked a final diagnosis due to presence of salmon-colored mucosa but no biopsies. Eight subjects in the Primary Analysis Population had BE, for a disease prevalence of 8.6%. Seven had short-segment BE (SSBE, <3 cm) and one had long-segment BE (LSBE, ≥3 cm). Of the eight subjects (8.6%) with BE on EGD, seven were EG positive; of the 85 (91.4%) subjects negative for BE, 69 were EG-negative, resulting in EG sensitivity of 87.5% (95% CI: 47.4%-99.7%) and specificity of 81.2% (95% CI: 71.2%-88.8%) (**Table 4**). Sensitivity for SSBE was 86% (95% CI: 42%-100%; n=7) and the sole case of LSBE was correctly detected by EG. PPV was 30.4% (95% CI: 13.2%-52.9%), and NPV was 98.6% (95% CI: 92.3%-99.96%). Overall agreement between EG and EGD results (i.e., test ‘accuracy’) was 81.7% (95% CI: 72.4%-89.0%).

**Table 2.** Summary of EsoCheck Cell Collection and EGD Procedure(s) (Main EsoCheck Population)

| **EsoCheck Cell Collection Procedure Summary** | **Total (n=180)**<br>**n (%)** |
| --- | --- |
| Was EsoCheck cell collection successfully performed? |  |
| No | 17 (9.4) |
| Yes | 163 (90.6) |
| Procedure duration in minutes |  |
| Mean | 5.8 |
| SD | 6.6 |
| Median | 4.0 |
| Min, max | 1, 70 |
| Were any adverse events or adverse device effect experienced? |  |
| No | 163 (90.6) |
| Yes | 0 (0.0) |
| **EGD Procedure Summary** | **Total (n=180)**<br>**n (%)** |
| Were EGD results collected? |  |
| No* | 17 (9.4) |
| Yes | 163 (90.6) |
| Salmon-colored mucosa (including tongues) present? |  |
| No | 123 (68.3) |
| Yes | 40 (22.2) |
| Length of salmon-colored mucosa |  |
| <1 cm | 12 (6.7) |
| ≥1 and < 3 cm | 23 (12.8) |
| ≥3 cm | 5 (2.8) |
| Were biopsies taken from the tubular esophagus?<sup>†</sup> |  |
| No | 1 (<1.0) |
| Yes | 39 (21.7) |

\*Subjects who failed EC cell collection were exited early from study participation and EGD results were not collected for performance endpoint analysis
<sup>†</sup>Subjects without salmon-colored mucosa on EGD were not required to undergo biopsy of the tubular esophagus
Percentages are based on the Main EsoCheck Population, N in header
Missing datapoints were not inferred

**Table 3.**
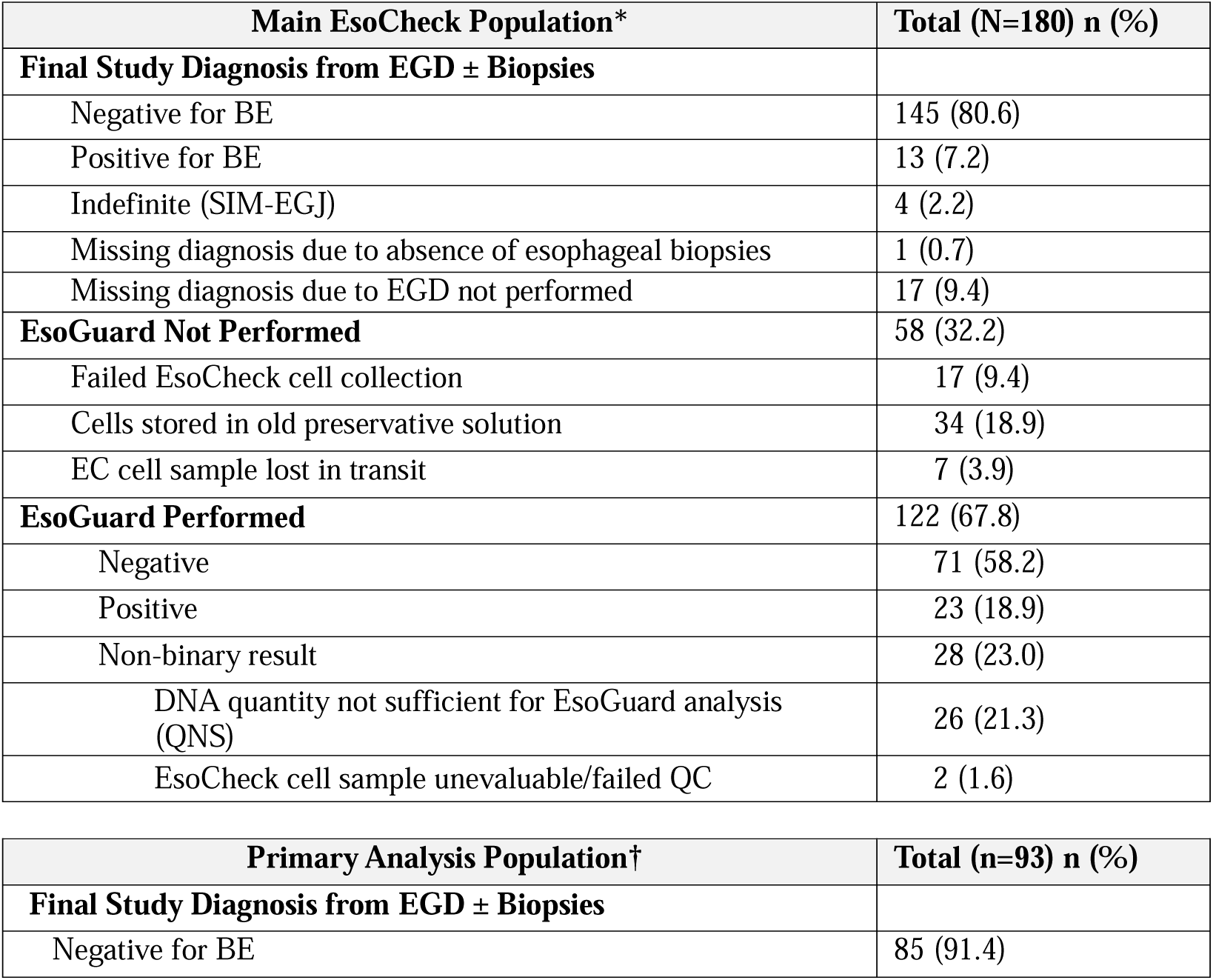
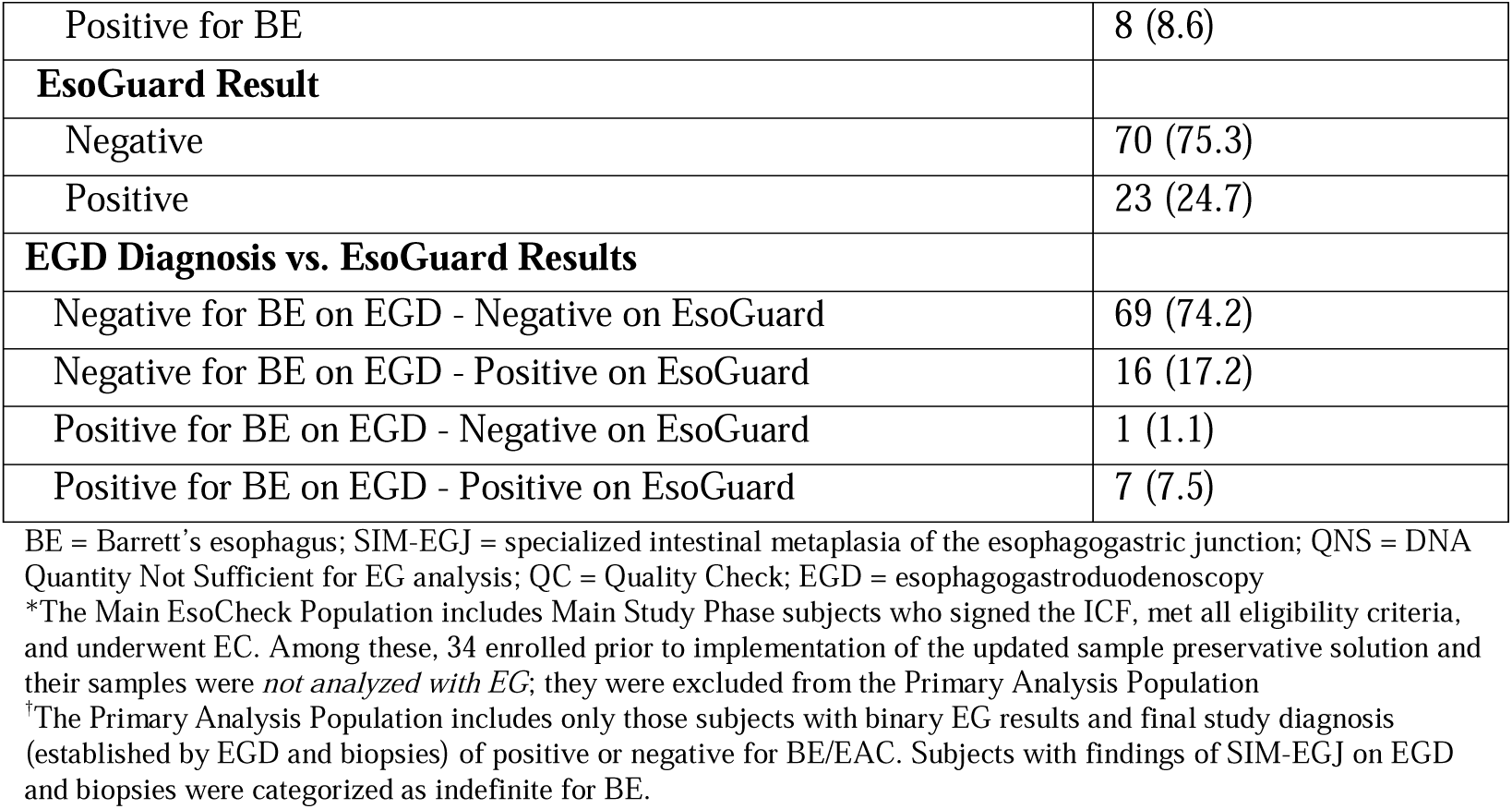
EGD and Biopsy-based Final Study Diagnosis, and EsoGuard Results.

| **Main EsoCheck Population\*** | **Total (N=180) n (%)** |
| --- | --- |
| **Final Study Diagnosis from EGD ± Biopsies** |  |
| Negative for BE | 145 (80.6) |
| Positive for BE | 13 (7.2) |
| Indefinite (SIM-EGJ) | 4 (2.2) |
| Missing diagnosis due to absence of esophageal biopsies | 1 (0.7) |
| Missing diagnosis due to EGD not performed | 17 (9.4) |
| **EsoGuard Not Performed** | 58 (32.2) |
| Failed EsoCheck cell collection | 17 (9.4) |
| Cells stored in old preservative solution | 34 (18.9) |
| EC cell sample lost in transit | 7 (3.9) |
| **EsoGuard Performed** | 122 (67.8) |
| Negative | 71 (58.2) |
| Positive | 23 (18.9) |
| Non-binary result | 28 (23.0) |
| DNA quantity not sufficient for EsoGuard analysis (QNS) | 26 (21.3) |
| EsoCheck cell sample unevaluable/failed QC | 2 (1.6) |

| **Primary Analysis Population†** | **Total (n=93) n (%)** |
| --- | --- |
| **Final Study Diagnosis from EGD ± Biopsies** |  |
| Negative for BE | 85 (91.4) |
| Positive for BE | 8 (8.6) |
| **EsoGuard Result** |  |
| Negative | 70 (75.3) |
| Positive | 23 (24.7) |
| **EGD Diagnosis vs. EsoGuard Results** |  |
| Negative for BE on EGD - Negative on EsoGuard | 69 (74.2) |
| Negative for BE on EGD - Positive on EsoGuard | 16 (17.2) |
| Positive for BE on EGD - Negative on EsoGuard | 1 (1.1) |
| Positive for BE on EGD - Positive on EsoGuard | 7 (7.5) |

BE = Barrett's esophagus; SIM-EGJ = specialized intestinal metaplasia of the esophagogastric junction; QNS = DNA Quantity Not Sufficient for EG analysis; QC = Quality Check; EGD = esophagogastroduodenoscopy
\*The Main EsoCheck Population includes Main Study Phase subjects who signed the ICF, met all eligibility criteria, and underwent EC. Among these, 34 enrolled prior to implementation of the updated sample preservative solution and their samples were *not analyzed with EG*; they were excluded from the Primary Analysis Population
†The Primary Analysis Population includes only those subjects with binary EG results and final study diagnosis (established by EGD and biopsies) of positive or negative for BE/EAC. Subjects with findings of SIM-EGJ on EGD and biopsies were categorized as indefinite for BE.

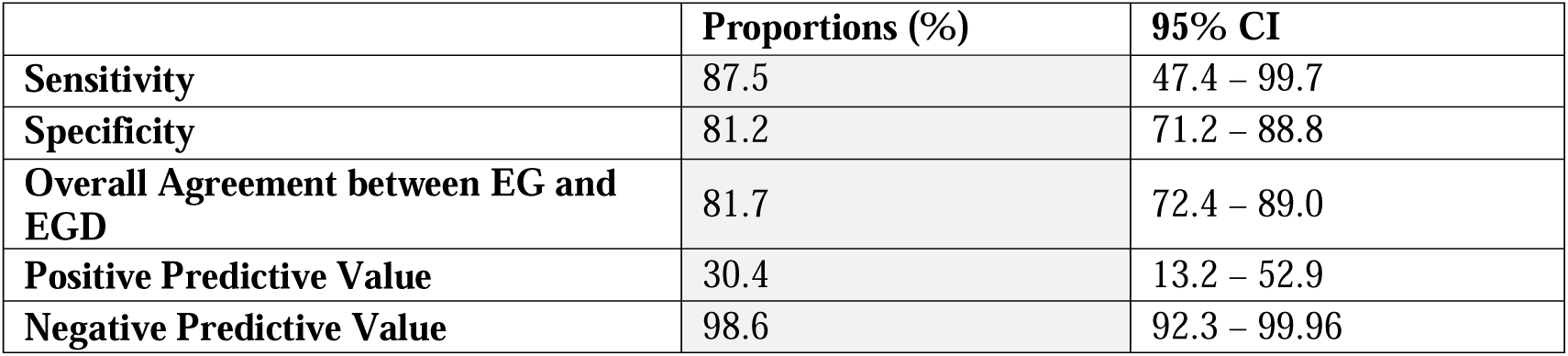

|  | **Proportions (%)** | **95% CI** |
| --- | --- | --- |
| **Sensitivity** | 87.5 | 47.4 – 99.7 |
| **Specificity** | 81.2 | 71.2 – 88.8 |
| **Overall Agreement between EG and EGD** | 81.7 | 72.4 – 89.0 |
| **Positive Predictive Value** | 30.4 | 13.2 – 52.9 |
| **Negative Predictive Value** | 98.6 | 92.3 – 99.96 |

EC acceptability from subject and device administrator surveys is in **Supplemental Table 3**. Most subjects expressed satisfaction with EC (>85%), and nearly 90% reported willingness to recommend it to friends or family needing BE screening. Over 87% of EC device administrators reported satisfaction with cell collection and 93% reported feeling comfortable performing EC.

One hundred thirty subjects underwent EGD on the same day as EC and were assessed for esophageal injury following EC (**Table 5**). Only two (2/130, 1.5%) esophageal abrasions were identified, both mild (superficial abrasions of the mucosa, < 5 mm in diameter, with no bleeding or oozing). Two other subjects experienced mild AEs deemed possibly related to EC: one scratchy throat and one episode of discomfort that resolved without intervention. Overall incidence of ADEs was 1.5% (4/265), with no serious AEs and no unanticipated ADEs.

**Table 5.** Summary of Adverse Device Effects (Safety Population)

| **Description of Event** | **Total (N=265) n (%\*)** |
| --- | --- |
| Scratchy throat | 1 (0.3) |
| Chest discomfort | 1 (0.3) |
| Esophageal abrasion assessed? |  |
| No, EsoCheck and EGD procedures performed on different dates | 33 (12.5) |
| No, EGD was not performed (DA Training Phase subjects) | 84 (31.7) |
| Yes | 130 (49.1) |
| **Esophageal abrasion present?** | **n = 130 n (%)** |
| No | 128 (98.5) |
| Yes | 2 (1.5) |
| **Abrasion Grade** | **n = 2 n (%)** |
| Grade 1 (Mild abrasions: < 5 mm in length with no visible oozing vessel) | 2 (100) |
| Grade 2 – 5 | 0 (0.0) |

## Discussion

We report the operating characteristics, safety, and tolerability of the first use of the EG/EC non-endoscopic screening technology in its intended-use population. Additionally, we describe alterations in the processing to facilitate transportation of the sample at ambient temperatures, an important improvement in its evolution as a point-of-service test. Our work demonstrates EG was safe and well-tolerated. Additionally, the alterations in processing do not seem to degrade performance of the assay compared to previous reports (although the number of BE cases in our population was not large).[22,23] While often not reported, implementation studies such as ours are vital to demonstrate the likely performance of the assay in a real-world setting. These data suggest that EG/EC screening should be suitable for use in high-risk patients to screen for BE.

Sensitivity of 87.5% and specificity of 81.2% was observed for EG detection of BE in this intended-use population, resulting in a PPV of 30.4%, NPV of 98.6%, and overall agreement between EG and EGD of 81.7%. This is consistent with previously published operating characteristics for EG/EC, the first of which was reported in a multi-center case-control study where the assay was performed by a single academic research laboratory on samples transported frozen.[20]. In that study EG sensitivity and specificity were 88% and 91.7%, respectively. A subsequent 243-subject multi-center case-control study was conducted by the National Cancer Institute (NCI)’s Barrett’s Esophagus Translational Research Network (BETRNet) where all samples were collected with EC, preserved and transported at room temperature, and EG performed by a commercial laboratory (LucidDx Labs).[21] Overall EG sensitivity and specificity for detecting BE and EAC was 85% and 85%, respectively.

EG/EC performance in our study was notable given that seven of the eight (7/8, 87.5%) EGD-positive patients had SSBE. This proportion of short segment disease is consistent with literature demonstrating SSBE accounts for ∼70% of disease in a screening population.[24, 25] Sensitivity of EG specifically for detection of SSBE was preserved at 86% in this study. In comparison, studies utilizing older sponge-on-a-string (SoS) cell collection devices reported SSBE sensitivity at 61.2-63%.[26, 27] While the current SSBE numbers are small, superior SSBE sensitivity in this and prior EG/EC studies may reflect both robustness of the EG assay and advances in non-endoscopic cell collection with the encapsulated balloon technology of EC. SoS devices collect cells indiscriminately from the entire esophagus and oropharynx, while the EC device allows anatomically targeted cell collection and sample protection during retrieval, which prevents dilution or contamination from cells outside the target area. SSBE sensitivity is critical for BE screening since most BE/EAC in the screening population is short segment disease.

Literature suggests that only 10-30% of chronic GERD patients undergo endoscopic BE screening, potentially due to limited knowledge of screening recommendations by primary care physicians, patient failure to report GERD symptoms, and hesitancy to undergo endoscopy.[14–16, 28, 29] Availability of a rapid, non-endoscopic, office-based test that can be performed by trained, non-physician providers should improve patient access to, and compliance with, BE screening. New in their 2022 updates, both the ACG and AGA recommended non-endoscopic cell collection technologies paired with a biomarker test as an acceptable alternative to EGD for initial screening of patients at elevated risk for BE/EAC.[11, 12]

Generally speaking, physicians seek to minimize false negatives and maximize sensitivity in screening tests. While the sensitivity and specificity of the EG/EC were 80-90% in our study, the PPV was 30.4%, due to the low prevalence of BE in our population (8.6%), which is consistent with the literature. While that means many EGDs performed in follow-up to a positive EG/EC might be expected to be negative for BE, the test is unlikely to miss cases of BE, and will unburden the system, because the three-fourths of patients with a negative result require no further work-up. The quarter of patients with positive results undergo an EGD that would have been recommended anyway, in the absence of a non-endoscopic, point-of-service test. Most importantly, a simple, point-of-service test such as this has the potential to “widen the top of the funnel” - to bring in a considerable proportion of patients who are currently denied the opportunity to be screened for this disease.

Important in any implementation study in the intended-use population, EC was safe and well-tolerated, with 90% of patients able to provide a sample. The median procedure time of four minutes offers the procedural efficiency needed for widespread screening. There were no serious adverse events, and esophageal abrasions of the mildest grade (superficial, <5 mm, no oozing/bleeding) occurred in only 1.5% of cases.

A key strength of this study is that it assessed EG performance in a screening (“intended use”) population of patients meeting national gastroenterology society BE screening guidelines. The prevalence of BE was consistent with the reported 5% to 15% prevalence in a real-world population with GERD and additional risk factors.[5, 30, 31] No cases of dysplasia or EAC were identified, consistent with the low incidence of these disease stages seen in larger, population-based studies.[32–34] These observations suggest our study population accurately reflects the real-world BE screening population and that the results, including the EG performance, are generalizable. Data from this study in a screening population supplements the two NCI-funded case-control studies previously mentioned,[20, 21] suggesting that its previously-reported operating characteristics will be preserved in the intended-use population. The fact that EG performance in this study is closely aligned with that reported in the two prior studies is encouraging, since case-control studies can overestimate the performance of a diagnostic test.[20, 21] Additionally, the study from the BETRNet consortium used the same laboratory as ours and demonstrated nearly identical EG sensitivity and specificity, providing further confidence in EG/EC as a BE screening tool.[21, 35]

There are several limitations to our study. During transport of specimens to LucidDx Labs, seven DNA samples were lost, reducing the number that were analyzed with EG. A second study limitation is the number of samples (26/122, 21.3%) with QNS. Most samples in this study (121/122, 99.2%), were extracted using an older, column-based DNA extraction method (QIAamp DNA Mini Kit, QIAGEN; Hilden, Germany). However, the DNA extraction method at LucidDx Labs has since been updated to a magnetic-bead based method (NucleoMag, Macherey-Nagel; Düren, Germany), which has reduced the QNS rate to ∼5%.[36, 37]. Therefore, we expect the QNS rate in future studies to be significantly lower than observed here. A third study limitation is the relatively small number of subjects with BE, resulting in a broad 95% confidence interval for sensitivity. Given that our reported sensitivity resembles other reports of EC/EG studies, we expect future studies evaluating EG in a screening population with larger sample sizes will yield similar sensitivities, but tighter confidence intervals. Finally, this study was limited to male subjects because the ACG guidelines prior to the 2022 update were equivocal in recommending BE screening for women. Current ACG guidelines *do* recommend screening for women with appropriate risk factors. The BETRNet study enrolled both sexes and found similar performance for overall disease detection in men and women.[21]

To conclude, this study in the intended-use screening population of patients at high-risk for BE/EAC demonstrated non-endoscopic cell collection with EsoCheck combined with EsoGuard DNA biomarker testing was well-tolerated, convenient, safe, and efficient, with acceptable accuracy for detecting disease. Our findings replicated results from previous case-control studies in terms of sensitivity and specificity for BE detection, with a PPV and a NPV that can appropriately guide clinical decision-making. EG/EC provides a potential point-of-service alternative to EGD for screening and detection of BE. Improved accessibility of non-endoscopic, in-office testing could increase BE screening rates, improve early disease detection, and ensure BE patients receive appropriate endoscopic surveillance and treatment to avoid progression to EAC.

## Data Availability

All data produced in the present study are available upon reasonable request to the authors

