## Supplemental Tables 1-3 for "Use of the EsoGuard^®^ Molecular Biomarker Test in Non-Endoscopic Detection of Barrett’s Esophagus among High-Risk Individuals in a Screening Population"

| **Supplemental Table 1. Inclusion and Exclusion Criteria** | |
| --- | --- |
| **Inclusion Criteria^[1]^** | **Exclusion Criteria** |
| 1. Male age 50 years or older; 2. ≥5 years of:  - GERD symptoms, - GERD treated with PPI therapy whether symptom control was achieved or not, or - Any combination of treated and untreated periods if the cumulative total is at least 5 years;  3. One or more of the following additional risk factors:  - White (Caucasian, non-Hispanic) race, - Current or history of tobacco/cigarette smoking, - Obesity as defined by a BMI of ≥30kg/m^2^, - First degree relative with BE or EAC;  4. No solid foods eaten for at least 2 hours prior to EsoCheck cell collection | 1. History of prior EGD procedure; 2. Inability to provide written informed consent; 3. On anticoagulant drug(s) that cannot be temporarily discontinued; 4. Known history of esophageal varices or esophageal stricture; 5. Any contraindication per the Investigator’s medical judgment for undergoing EC cell collection or EGD with biopsies, including but not limited to comorbidities such as coagulopathy, known history of esophageal diverticula, esophageal fistula and/or esophageal ulceration; 6. History (whether physician diagnosed, or patient reported) of dysphagia or odynophagia including with swallowing pills, sensation of food being stuck in the throat/esophagus, or difficulty tolerating dental procedures due to gagging; 7. Oropharyngeal tumor; 8. History of esophageal or gastric surgery, except for uncomplicated surgical fundoplication; 9. History of MI or CVA within the prior 6 months; 10. History of esophageal motility disorder; 11. Implanted Linx device |
| GERD – gastroesophageal reflux disease; PPI – proton pump inhibitor; BMI – body mass index; BE – Barrett’s Esophagus; EAC – esophageal adenocarcinoma; EGD – Esophagogastroduodenoscopy; MI – myocardial infarction; CVA – cerebrovascular accident  [1] The 2022 updated ACG guidelines on diagnosis and management of BE recommend screening in patients with chronic GERD and 3 or more of the following risk factors: male sex, age>50 years, white race, history of tobacco smoking, obesity, and family history of BE or EAC in a 1^st^ degree relative | |

| **Supplemental Table 2. Esophageal Abrasion Grading System** | |
| --- | --- |
| **Abrasion Grade** | **Description** |
| Grade 1 | **Mild abrasions:** one or more abrasions <5 mm in length with no visible oozing vessel |
| Grade 2 | **Mild abrasions:** one or more abrasions ≥5 mm in length with no visible oozing vessel |
| Grade 3 | **Moderate abrasions**: one or more abrasions with no active bleeding but minimal ooze from vessel without compromising mucosal views during endoscopy |
| Grade 4 | **Severe abrasions:** one or more abrasions with no active bleeding but with some oozing blood which compromises mucosal views during endoscopy |
| Grade 5 | **Complicated abrasions:** one or more abrasions resulting in active bleeding and requiring endoscopic or surgical intervention |
| **Source:** Januszewicz, W, Tan WK, Lehovsky K, et al. Safety and acceptability of esophageal Cytosponge cell collection device in a pooled analysis of data from individual patients. Clin Gastroenterol Hepatol, 2019;17:647–56. | |

Supplemental Table 3. Summary of User Acceptability (Main Phase Subjects who Underwent EsoCheck)

| **User Acceptability summary** | **Total (n=181) n (%*)** |
| --- | --- |
| **Pre-Procedure Subject Survey:** |  |
| I feel relief knowing I will be screened for esophageal pre-cancer/cancer. |  |
| Strongly agree | 137 (76.7) |
| Somewhat agree | 26 (14.4) |
| Neither agree nor disagree | 15 (8.3) |
| Somewhat disagree | 0 (0.0) |
| Strongly disagree | 0 (0.0) |
| I feel confident that I will be able to tolerate the procedure today. |  |
| Strongly agree | 89 (49.2) |
| Somewhat agree | 56 (30.9) |
| Neither agree nor disagree | 28 (15.5) |
| Somewhat disagree | 2 (1.1) |
| Strongly disagree | 3 (1.7) |
| I am anxious about having the procedure today. |  |
| Strongly agree | 28 (15.5) |
| Somewhat agree | 58 (32.0) |
| Neither agree nor disagree | 28 (15.5) |
| Somewhat disagree | 21 (11.6) |
| Strongly disagree | 43 (23.8) |
| **Post-Procedure Subject Survey:** |  |
| The procedure went better than I expected. |  |
| Strongly agree | 90 (49.7) |
| Somewhat agree | 39 (21.6) |
| Neither agree nor disagree | 23 (12.7) |
| Somewhat disagree | 13 (7.2) |
| Strongly disagree | 13 (7.2) |
| I would be willing to undergo this procedure again if needed. |  |
| Strongly agree | 118 (65.2) |
| Somewhat agree | 35 (19.3) |
| Neither agree nor disagree | 12 (6.6) |
| Somewhat disagree | 2 (1.1) |
| Strongly disagree | 11 (6.1) |
| I was satisfied with how the procedure went. |  |
| Strongly agree | 129 (71.3) |
| Somewhat agree | 27 (14.9) |
| Neither agree nor disagree | 9 (5.0) |
| Somewhat disagree | 4 (2.2) |
| Strongly disagree | 9 (5.0) |
| I would recommend this procedure to a family member or friend in need of esophageal pre-cancer/cancer screening. |  |
| Strongly agree | 127 (70.2) |
| Somewhat agree | 33 (18.2) |
| Neither agree nor disagree | 7 (3.9) |
| Somewhat disagree | 3 (1.7) |
| Strongly disagree | 8 (4.4) |
| **Post-Procedure EsoCheck Device Administrator/Investigator Survey:** |  |
| The patient seemed anxious about the procedure. |  |
| Strongly agree | 21 (11.6) |
| Somewhat agree | 47 (26.0) |
| Neither agree nor disagree | 8 (4.4) |
| Somewhat disagree | 31 (17.1) |
| Strongly disagree | 71 (39.2) |
| The patient experienced discomfort during the procedure. |  |
| Strongly agree | 11 (6.1) |
| Somewhat agree | 59 (32.6) |
| Neither agree nor disagree | 18 (9.9) |
| Somewhat disagree | 35 (19.3) |
| Strongly disagree | 55 (30.4) |
| The patient was able to tolerate the procedure. |  |
| Strongly agree | 115 (63.5) |
| Somewhat agree | 41 (22.7) |
| Neither agree nor disagree | 8 (4.4) |
| Somewhat disagree | 5 (2.8) |
| Strongly disagree | 9 (5.0) |
| I was satisfied with how the procedure went. |  |
| Strongly agree | 130 (71.8) |
| Somewhat agree | 27 (14.9) |
| Neither agree nor disagree | 8 (4.4) |
| Somewhat disagree | 3 (1.7) |
| Strongly disagree | 10 (5.5) |
| I felt comfortable performing the procedure in this patient. |  |
| Strongly agree | 135 (75) |
| Somewhat agree | 33 (18) |
| Neither agree nor disagree | 3 (2) |
| Somewhat disagree | 5 (3) |
| Strongly disagree | 2 (1) |
| **Telephone Contact Survey Questions:** |  |
| I am glad I underwent the EsoCheck procedure. |  |
| Strongly agree | 121 (66.9) |
| Somewhat agree | 35 (19.3) |
| Neither agree nor disagree | 6 (3.3) |
| Somewhat disagree | 1 (<1.0) |
| Strongly disagree | 3 (1.7) |
| I prefer this procedure to EGD. |  |
| Strongly agree | 69 (38.1) |
| Somewhat agree | 23 (12.7) |
| Neither agree nor disagree | 24 (13.3) |
| Somewhat disagree | 20 (11.1) |
| Strongly disagree | 30 (16.6) |

EGD – Esophagogastroduodenoscopy

*Percentages may not total 100 due to rounding; percentages are based on the Main Study Phase subjects who underwent EsoCheck (whether successfully or unsuccessfully), N in the header
